# Cutting Through the Noise: Stochastic Pulse Timing for Deep Brain Stimulation

**DOI:** 10.64898/2026.07.08.26357382

**Authors:** Matthew R Baker, Hemant Bokil, Soroush Niketeghad, Kai J Miller, Bryan T Klassen

**Affiliations:** Department of Neurosurgery, Mayo Clinic; Boston Scientific Corporation; Department of Biomedical Engineering and Physiology, Mayo Clinic; Department of Neurology, Mayo Clinic

**Keywords:** Deep Brain Stimulation, Stochastic Pulse Timing, Dysarthria, Essential Tremor

## Abstract

**Background:** Deep brain stimulation (DBS) is a widely used therapy for neurologic and psychiatric disorders. Conventional DBS delivers highly regular stimulation patterns that suppress pathological activity but can induce stimulation-related side effects, limiting the therapeutic window. Introducing controlled temporal variability through stochastic pulse timing may represent an alternative programming dimension to improve tolerability while preserving clinical benefit.

**Methods:** An adult in their 60’s with bilateral Vim DBS underwent evaluation of tonic, pink-noise, and white-noise stimulation patterns delivered through his chronically implanted Boston Scientific Genus system using the Chronos research platform. We assessed tremor and stimulation-induced side effects using accelerometry, spiral drawing tasks, standardized speech recordings, and patient-reported paresthesias.

**Results:** Pink noise stimulation preserved meaningful tremor suppression while improving tolerability compared with conventional tonic 130 Hz stimulation. Under tonic stimulation, dysarthria and paresthesias were prominent at 2.0 mA, narrowing the usable therapeutic window. In contrast, pink noise maintained tremor control across the same amplitude range with reduced side-effect burden. White noise stimulation demonstrated intermediate effects, providing improved tolerability relative to tonic stimulation but less tremor suppression than pink noise. Findings were consistent across accelerometry and functional drawing tasks.

**Conclusion:** This study provides first-in-human evidence that temporally structured stochastic pulse timing can preserve therapeutic benefit while expanding the tolerable stimulation range relative to tonic DBS. These findings suggest that temporal structure represents a clinically meaningful programming dimension that may broaden the DBS therapeutic window using software based updates to existing hardware. Further evaluation in larger cohorts is warranted.

## Introduction

Deep brain stimulation (DBS) is an established therapy for neurologic and psychiatric disorders and is thought to work by suppressing pathological brain network activity [1–3]. Most current DBS paradigms rely on tonic high-frequency stimulation (130-185Hz) with fixed inter-pulse intervals (IPIs). While clinically effective, these highly regular pulses may not be the most efficient way to address pathologically synchronized network activity and may result in stimulation-induced side effects. [4–6]. Whether introducing structured variation into pulse timing could offer a more effective or better-tolerated alternative to tonic stimulation has not been well-explored.

In essential tremor, DBS targeting the ventral intermediate nucleus (Vim) of the thalamus provides rapid and robust tremor relief [7, 8] (Fig 1A). Yet, treatment efficacy is frequently constrained by stimulation-induced dysarthria and paresthesias, reported in up to 50% of patients [9–11](Fig 1B). These side effects often force patients and clinicians to choose between incomplete tremor control and tolerability, leading to suboptimal outcomes and frequent reprogramming.

**Fig. 1.**
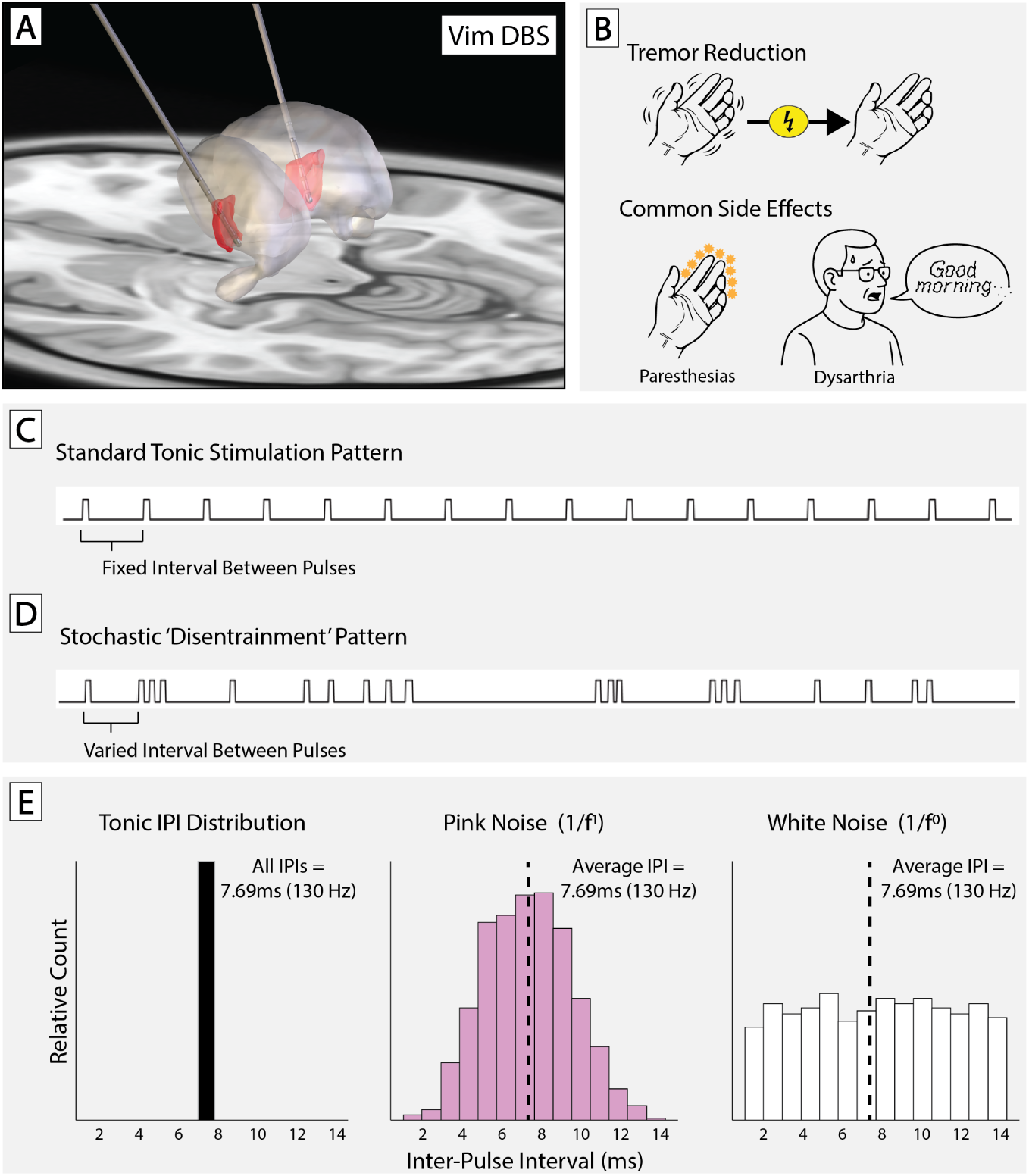
Stochastic ‘Disentrainment’ Stimulation Patterns for Deep Brain Stimulation (DBS). **(A)** Schematic of DBS targeting the ventral intermediate nucleus (Vim) of the thalamus for essential tremor. **(B)**While effective for tremor control, conventional tonic high frequency DBS can cause stimulation-induced side effects such as paresthesias and dysarthria. **(C)** Standard tonic stimulation delivers pulses at fixed intervals (constant inter-pulse interval, IPI). **(D)** Stochastic “disentrainment” patterns introduce variability in the IPI while maintaining the same average frequency. **(E)** Distributions of IPIs across patterns: tonic stimulation has a fixed IPI of 7.69ms (130Hz); pink noise follows a 1/f distribution centered at 7.69ms; white noise uses uniform random variation around the same mean IPI.

Temporally patterned stimulation introduces variability into the timing between pulses while preserving the average stimulation frequency [12, 13]. Noise patterns with distinct temporal structure—such as white noise, pink noise, and other 1/f-type processes—offer different ways of introducing correlated temporal variability across timescales. Many neural systems naturally exhibit 1/f-like temporal structure [14]. For example, pink noise follows a 1/f distribution and embeds scale-free temporal correlations, whereas white noise produces uncorrelated, randomly varying inter-pulse intervals. Such controlled variability may modulate circuits differently than tonic stimulation and achieve more favorable symptom and side-effect outcomes. Until recently, however, delivering these complex patterns in chronically implanted humans was not technically feasible.

We explored this question using the Boston Scientific Chronos research platform, which enables delivery of custom temporal stimulation patterns. Using this framework, we delivered custom pink-noise and white-noise stimulation patterns and evaluated their effects in a patient with essential tremor. Across electrode configurations and multiple amplitude titrations, noise-patterned stimulation consistently provided therapeutically meaningful tremor suppression while markedly improving tolerability, reducing stimulation-induced dysarthria and paresthesias across a broad range of settings. These findings provide the first in-human evidence that temporally structured pulse timing may meaningfully expand the therapeutic window of DBS.

## Methods

### Ethics Statement

The patient gave written consent to participate in a research protocol involving stochastic DBS patterns. All study procedures and the consent process were approved by Mayo Clinic’s internal review board (IRB no. 19-009878).

### Subject

The participant was a right-handed adult in their 60’s with a long-standing history of medication-refractory essential tremor, characterized by bilateral upper extremity tremor (left greater than right) that impaired fine motor tasks and daily functioning. He had previously undergone bilateral implantation of Boston Scientific Vercise Cartesia directional DBS leads, connected to a rechargeable Genus IPG system targeting the Vim nucleus of the thalamus.

While initial programming achieved substantial tremor reduction, the therapeutic window was limited by stimulation-induced dysarthria, which emerged at amplitudes required for full symptom control (Program 1). These side effects proved functionally limiting and prompted clinical reprogramming efforts. Although a revised directionally-steered multipolar configuration reduced the severity of dysarthria (Program 2), the patient remained unable to achieve complete tremor suppression without intolerable adverse effects. Contact configurations are presented in Figure S1.

### Stimulation Platform and Temporal Pattern Design

Stimulation was delivered through the patient’s chronically implanted Boston Scientific (Boston Scientific Corporation, Malborough, MA, USA) Genus deep brain stimulation system using the Chronos research platform, which enables real-time, pulse-by-pulse control of stimulation timing. This interface allows user-defined IPI sequences to be executed directly on the implanted pulse generator without hardware modification, enabling systematic manipulation of temporal pulse structure while maintaining commercially approved amplitude and charge density limits.

The conceptual framework for temporally patterned stimulation is illustrated in Fig 1. Conventional tonic deep brain stimulation delivers pulses at fixed intervals, resulting in highly regular temporal structure (Fig 1C). In contrast, stochastic stimulation paradigms introduce controlled variability into inter-pulse intervals while preserving the same average frequency and pulse width (Fig 1D–E). In this study, temporal structure was treated as the primary independent variable, with mean stimulation frequency (130 Hz) and pulse width (60 *µ*s) held constant across conditions.

Three stimulation paradigms were implemented: (1) tonic stimulation with fixed interpulse interval (7.69 ms), (2) pink noise–patterned stimulation in which IPIs followed a 1/f distribution introducing structured, scale-free temporal correlations, and (3) white noise–patterned stimulation in which IPIs varied randomly with a flat spectral profile and no temporal correlations. For stochastic paradigms, approximately five seconds of IPIs were generated according to the target distribution and cyclically repeated to produce continuous stimulation trains (due to hardware constraints).

Representative temporal statistics for each paradigm are shown in Fig S2. Tonic stimulation demonstrates constant IPIs with flat spectral density and negligible autocorrelation beyond lag zero. Pink noise stimulation exhibits a power-law spectral slope and long-range temporal correlations in the IPI sequence, whereas white noise stimulation shows flat spectral density with minimal autocorrelation. These analyses confirm that stimulation conditions differed only in temporal organization while maintaining equivalent mean pulse rate.

In addition to comparing temporal pulse-timing patterns, we evaluated pulse recharge configuration for tonic 130 Hz conditions. DBS pulses are charge-balanced by a recharge phase that can be delivered using either passive or active recharge. In passive recharge, charge dissipates naturally through the tissue–electrode interface, whereas active recharge applies a controlled anodic current to restore charge balance more rapidly. To determine whether the improved tolerability observed with stochastic stimulation was attributable to recharge characteristics rather than pulse timing, conventional tonic 130 Hz stimulation was tested using both active and passive recharge at identical amplitudes and pulse widths. This comparison isolated the effects of recharge mode from those of temporally patterned inter-pulse interval variability.

### Experimental Testing Blocks

Testing was organized into discrete experimental blocks designed to evaluate the effects of temporal stimulation structure across multiple behavioral paradigms and stimulation configurations. These testing blocks were conducted across three in-clinic DBS programming sessions and blinded home trials. Following the first two in-clinic sessions, the participant completed blinded one-week home trials using selected tonic and stochastic stimulation programs during routine daily activities. At the end of each trial, the patient provided qualitative feedback regarding tremor control, speech clarity, and overall program preference.

#### Accelerometry-based tremor assessment

Objective tremor suppression and side-effect profiles were evaluated using accelerometry during postural pen-hold and finger-to-nose tasks. Testing included five stimulation conditions: stimulation OFF, tonic 130 Hz stimulation at 1.0 and 2.0 mA, and pink noise–patterned stimulation at 2.0 and 3.0 mA delivered using Program 1 contact configuration (Fig S1). Outcome measures included accelerometer-derived tremor amplitude, dysarthria assessment during standardized speech tasks, and paresthesia ratings (Fig 2-4).

**Fig. 2.**
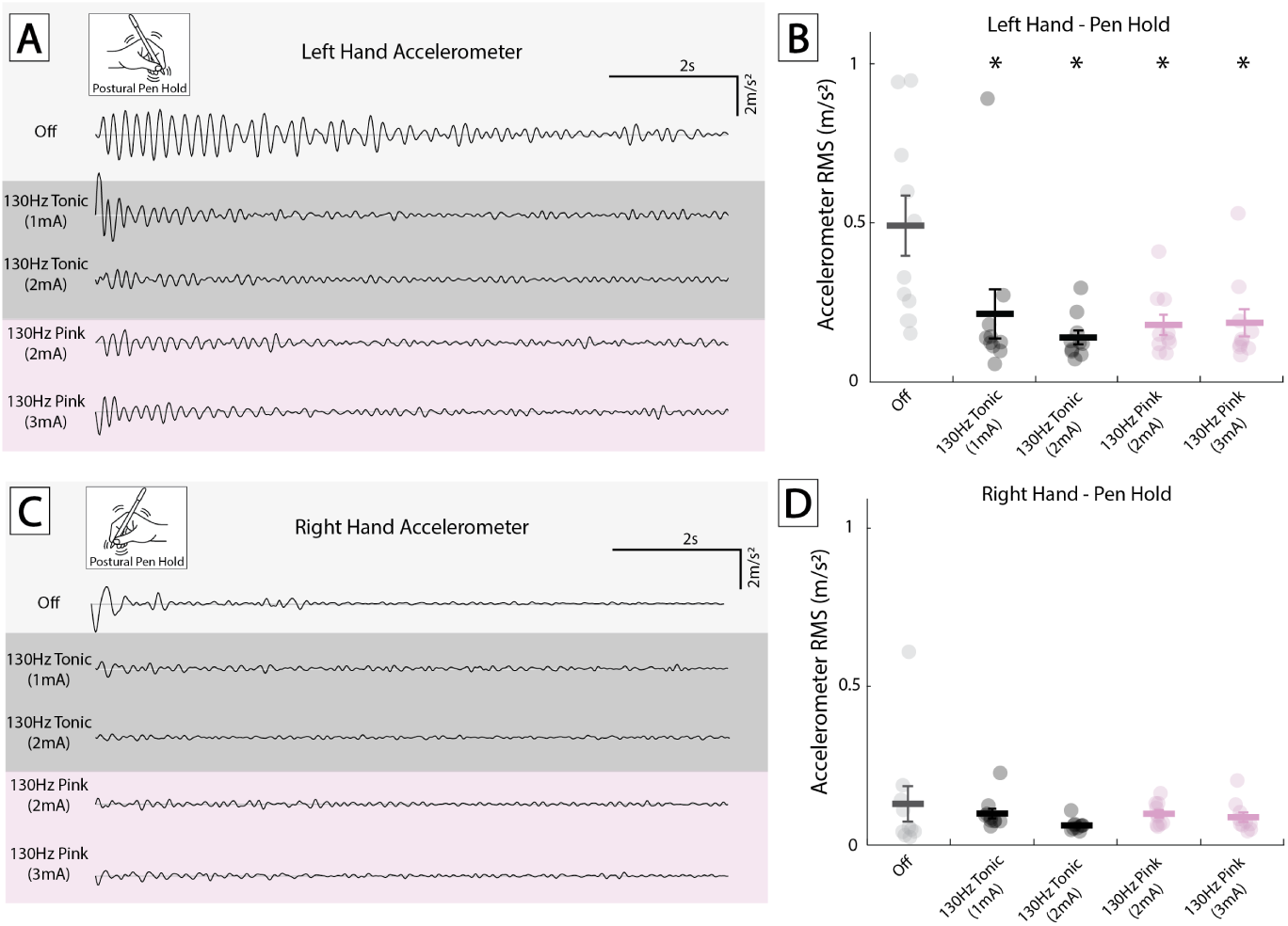
Postural tremor during different stimulation conditions at first clinic visit. **(A)** Representative accelerometer traces from the left hand during a postural pen-hold task under five stimulation conditions: OFF, tonic 130Hz at 1mA and 2mA, and pink noise-patterned stimulation at 2mA and 3mA. Both tonic and pink noise stimulation reduced tremor amplitude relative to baseline. **(B)** Quantification of 1 second bins for left-hand tremor using root mean square (RMS) amplitude of the filtered signal (first principal component, 4–12 Hz band) in 1 second bins. Asterisks indicate stimulation conditions significantly different from OFF (∗*p <* 0.05, paired Wilcoxon signed-rank tests with Holm correction following Friedman test) **(C)** Right-hand acceleromter traces during the same task and conditions. **(D)** RMS tremor amplitude for the right hand in 1 second bins. Baseline tremor was lower on the right, with no significant differences between conditions. All stimulation conditions used the program 1 contact configuration (monopolar; Fig S1).

**Fig. 3.**
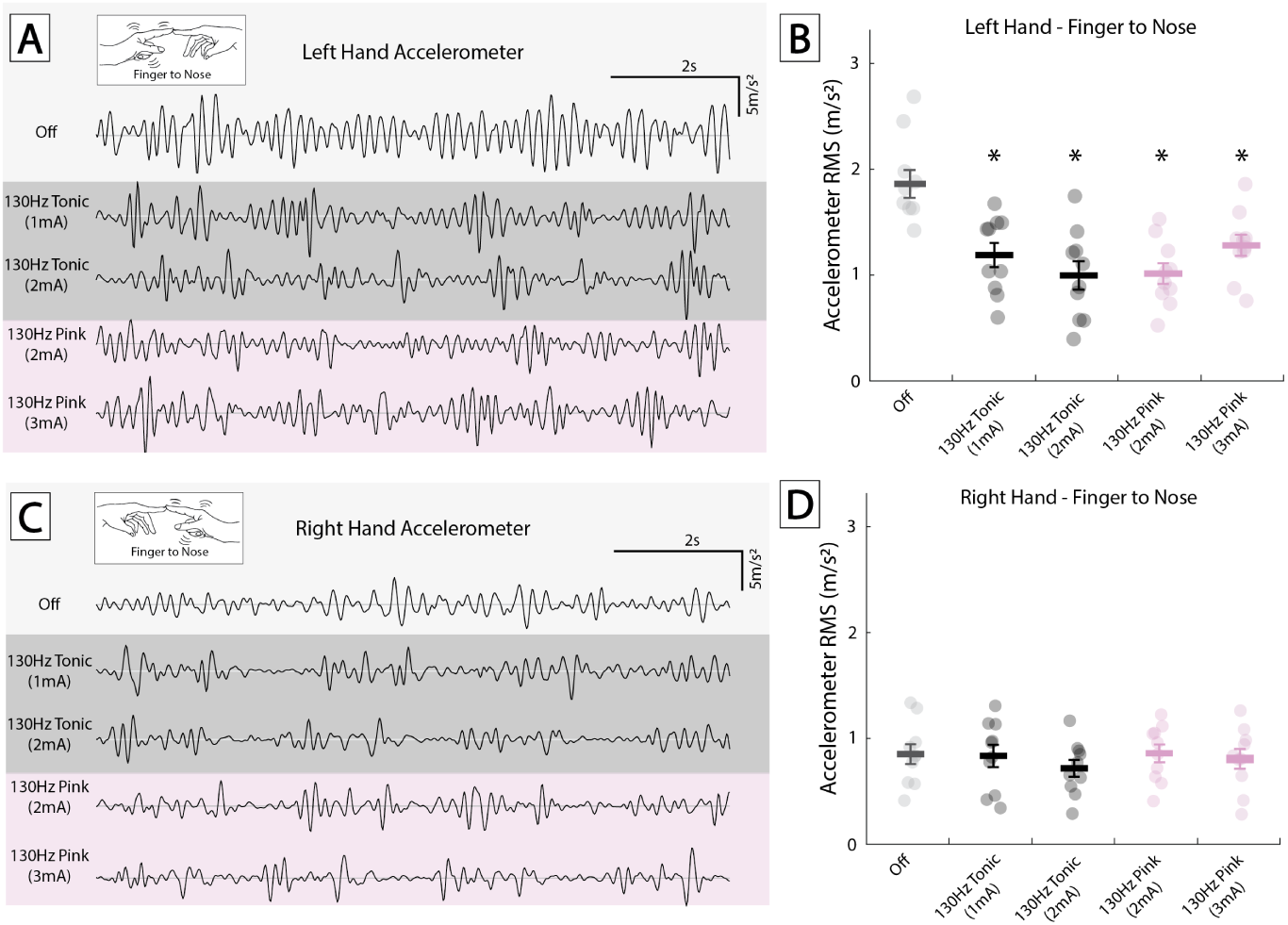
Kinetic tremor during finger-to-nose task at first clinic visit. **(A)** Accelerometer traces from the left hand during a finger-to-nose task under five stimulation conditions: OFF, tonic 130 Hz at 1 mA and 2 mA, and pink noise–patterned stimulation at 2 mA and 3 mA. **(B)** Quantification of 1 second bins for left-hand tremor using root mean square (RMS) amplitude of the filtered accelerometer signal (first principal component, 4–12 Hz band). Asterisks indicate stimulation conditions significantly different from OFF (∗*p <* 0.05, paired Wilcoxon signed-rank tests with Holm correction following Friedman test). **(C)** Right hand accelerometer traces showed lower baseline tremor, with minimal changes across stimulation conditions. **(D)**RMS amplitude for the right hand confirmed little difference from baseline. Tremor suppression was more evident in the left hand, which had higher initial tremor severity. All stimulation conditions used the program 1 contact configuration (monopolar; Fig S1).

**Fig. 4.**
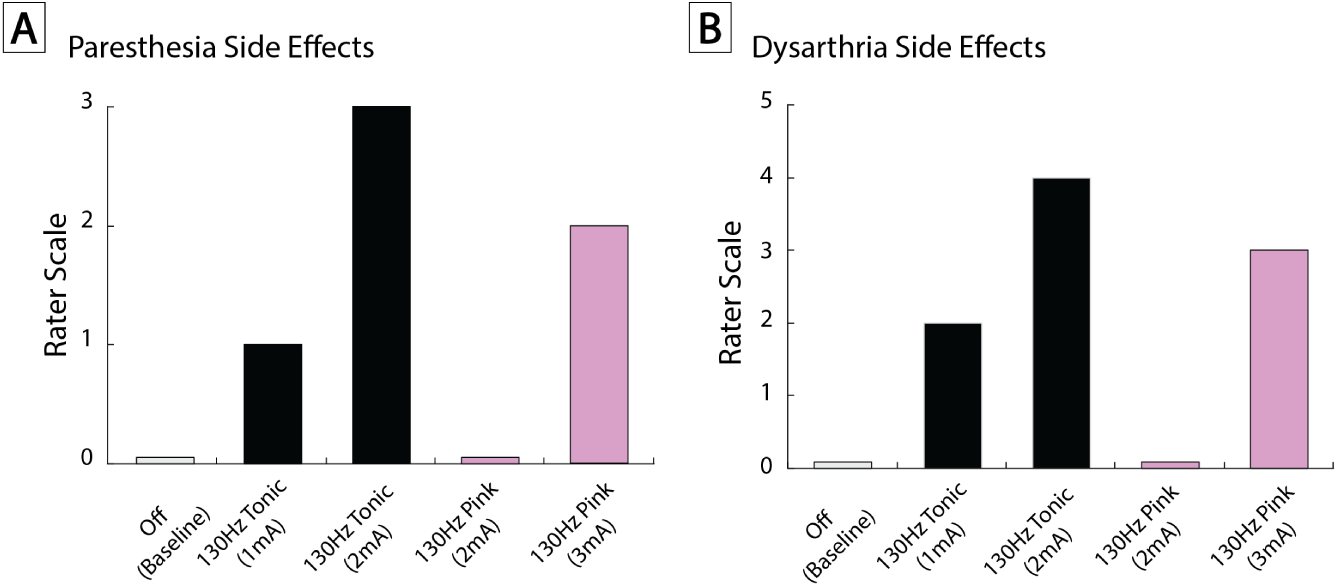
Stimulation-induced side effects at first clinic visit: paresthesias and dysarthria. **(A)** Paresthesia severity was rated by the patient using a 0–3 ordinal scale (0 = none, 3 = severe and bothersome). Tonic stimulation at 130 Hz and 2 mA produced the most bothersome paresthesias (score = 3), whereas pink noise stimulation at the same amplitude resulted in no reported paresthesias (score = 0). At 3 mA, pink noise stimulation caused mild paresthesias (score = 2). **(B)** Dysarthria was rated by a blinded movement disorders neurologist using a 0–5 scale (0 = no dysarthria, 5 = severe dysarthria). The most severe dysarthria occurred during 130 Hz tonic stimulation at 2 mA (score = 4), whereas pink noise stimulation at the same amplitude eliminated dysarthria (score = 0). The 3 mA pink noise condition was associated with mild dysarthria (score = 3). These findings demonstrate that pink noise stimulation preserved tolerability at amplitudes that produced intolerable side effects with conventional tonic DBS. All stimulation conditions used the program 1 contact configuration (monopolar; Fig S1).

#### Drawing-based amplitude testing

To more precisely characterize amplitude-dependent effects and therapeutic window, spiral drawing tasks were performed across expanded amplitude ranges (1.0, 2.0, 3.0, 4.0 mA) using tonic and pink noise-patterned stimulation. Testing was conducted using program 2 contact configuration (Fig S1), with amplitudes incrementally increased up to 4mA to assess tremor suppression alongside dysarthria and paresthesia side effects. Tremor severity was quantified using the Fahn-Tolosa-Marin (FTM) tremor scale (Fig 5, S3).

**Fig. 5.**
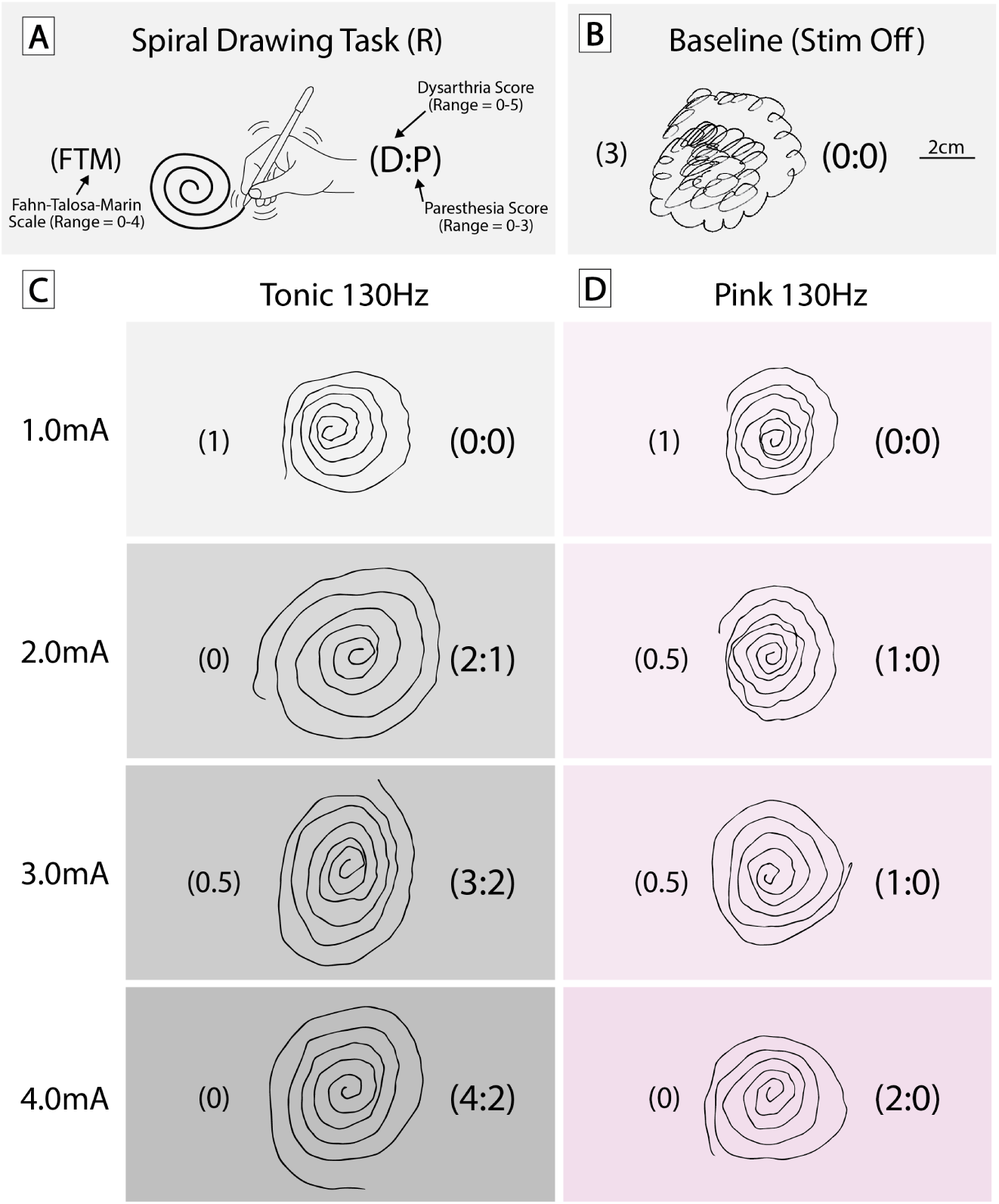
Right-hand spiral drawings and side effect profiles during in-clinic stimulation testing comparing tonic and pink noise stimulation. **(A)** Spiral drawing task schematic illustrating the Fahn–Tolosa–Marin (FTM) tremor scale (0–4), dysarthria score (0–5), and paresthesia score (0–3). **(B)** Baseline spiral drawing with stimulation OFF. Right-hand spiral drawings obtained during in-clinic testing under tonic 130 Hz **(C)** and pink noise–patterned 130 Hz **(D)** stimulation at 1.0, 2.0, 3.0, and 4.0 mA. FTM tremor scores are shown left of each spiral, and dysarthria and paresthesia scores are displayed in parentheses to the right (dysarthria:paresthesia). Both stimulation paradigms reduced tremor relative to OFF across amplitudes. At higher amplitudes, tonic 130 Hz was associated with increasing dysarthria and/or paresthesia, whereas pink 130 Hz achieved comparable tremor suppression with lower side effect scores. All stimulation conditions were tested using program 2 contact configurations (Fig S1).

To determine whether clinical effects observed with temporally patterned stimulation could be attributed to recharge dynamics rather than temporal structure, tonic 130Hz stimulation was tested under both passive and active recharge configurations across matched amplitudes (Fig S4-S5).

#### Noise structure

To assess whether any effects were due to the existence of temporal variability or its statistical structure, pink noise-patterned stimulation was directly compared with white noise-patterned stimulation across matched amplitudes (1.0, 1.5, 2.0 mA) using spiral and line drawing tasks. Tremor severity (FTM) and dysarthria/paresthesia side effects were assessed under each condition (Fig 6, S6).

**Fig. 6.**
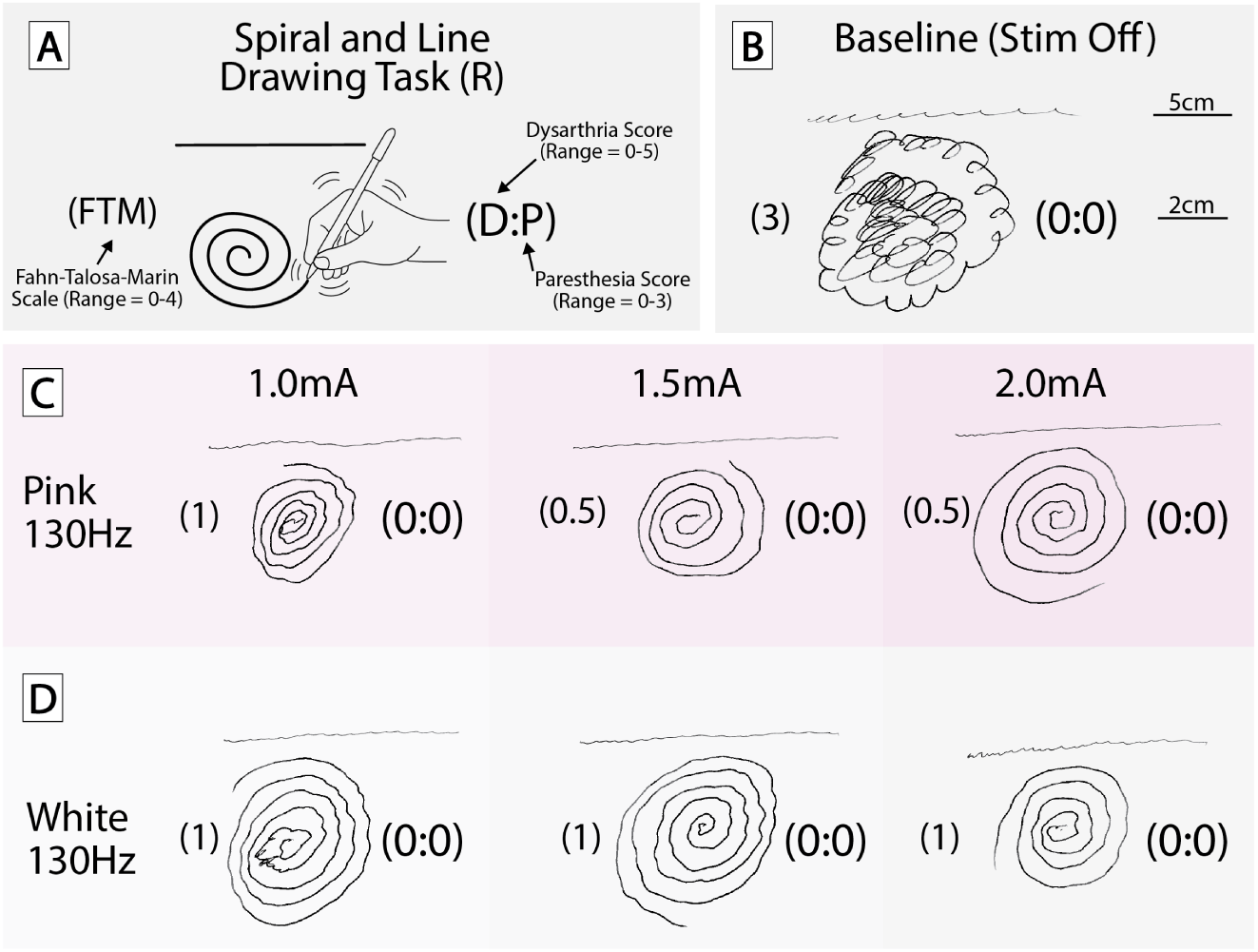
Right-hand spiral and line drawings during in-clinic stimulation testing comparing pink and white noise stimulation. **(A)** Spiral and line drawing task schematic illustrating the Fahn–Tolosa–Marin (FTM) tremor scale (0–4), incorporating both spiral and line components, along with dysarthria score (0–5) and paresthesia score (0–3). **(B)** Baseline drawing with stimulation OFF. Representative right-hand drawings obtained during in-clinic testing under pink noise–patterned 130 Hz **(C)** and white noise–patterned 130 Hz **(D)** stimulation at 1.0, 1.5, and 2.0 mA. FTM tremor scores are shown to the left of each drawing, and dysarthria and paresthesia scores are displayed in parentheses to the right (dysarthria:paresthesia). Both stimulation paradigms reduced tremor severity relative to OFF, with pink noise stimulation demonstrating slightly lower FTM scores at intermediate amplitudes compared with white noise. All stimulation conditions were tested using program 2 contact configurations (Fig S1).

### Outcome Measures

Tremor severity was assessed using both accelerometry-based metrics and drawing-based clinical scoring depending on the experimental paradigm. Drawing-based assessments included spiral and line drawing tasks. Tremor severity was rated using the FTM tremor rating scale (0–4) by a blinded board-certified movement disorders neurologist (BTK). On this scale, 0 = no tremor, 1 = mild tremor without functional impairment, 2 = moderate tremor with visible oscillation but preserved task completion, 3 = marked tremor with impaired drawing quality, and 4 = severe tremor with substantial distortion or inability to adequately complete the task [15].

Speech performance was evaluated using a brief, standardized reading passage (first 3 sentences of the Rainbow Passage [16]), read aloud during each stimulation condition. Audio recordings were reviewed offline (BTK), blinded to condition. Dysarthria was rated on a 0–5 ordinal scale, with 0 indicating normal speech and 5 indicating severe dysarthria impairing intelligibility.

Paresthesias and other stimulation-induced side effects were assessed through structured self-report immediately after each condition. The patient rated paresthesia severity on a 0–3 scale: 0 = none, 1 = mild/transient, 2 = mild but persistent, and 3 = moderate to severe and bothersome.

### Signal processing and statistics

Accelerometry data were analyzed using custom MATLAB scripts (MathWorks, Natick, MA). Signals from tri-axial sensors were bandpass filtered between 4–12 Hz to isolate tremor-related oscillations. The dominant tremor axis was identified using principal component analysis (PCA), and the first principal component was used for quantification. Tremor amplitude was calculated as the root mean square (RMS) of the filtered signal. For postural tasks, RMS values were computed in consecutive 1-second bins across the recording period. For kinetic tasks, segments corresponding to active movement during finger-to-nose testing were identified prior to RMS calculation.

Given the within-subject repeated-measures design and non-normal distribution of tremor amplitude values, nonparametric tests were used for accelerometry data. Overall differences across stimulation conditions were assessed using Friedman tests. When significant, post hoc comparisons versus the OFF condition were performed using paired Wilcoxon signed-rank tests with Holm correction for multiple comparisons. Statistical significance was defined as *p <* 0.05. The remainder of analyses were descriptive in nature given the single-subject design.

## Results

### Accelerometry-based tremor assessment

Tremor suppression and side-effect profiles were evaluated using accelerometry during postural pen-hold and finger-to-nose tasks across five stimulation conditions (OFF; tonic 130 Hz at 1.0 and 2.0 mA; pink noise–patterned stimulation at 2.0 and 3.0 mA). As shown in Figure 2A, the left-hand pen-hold condition demonstrated prominent postural tremor in the off state, which was substantially reduced during both tonic and pink-noise stimulation. Quantifying tremor stability across ten consecutive one-second bins revealed significantly lower RMS amplitude relative to off for all active stimulation conditions (Friedman *χ*^2^(4)=19.84, p=0.0005; all Holm-corrected p *<* 0.03)(Fig 2B). For the right hand during pen-hold, raw traces show some transient tremor suppression in the initial seconds of stimulation (Figure 2C). However, overall tremor amplitude was minimal for this task, and no stimulation condition differed significantly from the off state (all p *>* 0.05; Fig 2D).

During the finger-to-nose task, kinetic tremor was substantially larger than in the postural task, particularly in the left hand (Fig 3A). RMS tremor amplitude during the off condition exceeded 1.8 m/s², and all active stimulation conditions significantly reduced tremor (Friedman *χ*²(4)=19.52, p=0.0006; all Holm-corrected p *≤* 0.02)(Fig 3B). For the right hand during finger-to-nose, kinetic tremor was minimal and there were no significant differences between stimulation conditions (Fig 3D). While 130 Hz tonic and pink noise stimulation both produced clinically meaningful reduction in tremor amplitude across tasks, the tonic stimulation was also associated with prominent stimulation-induced side effects, limiting its clinical utility.

Side effects were rated on an ordinal clinical scale assessing paresthesias (range: 0-3) and dysarthria (range: 0-5; Fig 4). Paresthesias increased with tonic stimulation amplitude, reaching moderate severity at 2 mA (rating = 3), while pink-noise stimulation at the same amplitude produced no sensory disturbance (rating = 0). A higher-amplitude pink-noise condition (3 mA) produced only mild paresthesias (rating = 2), remaining more tolerable than tonic stimulation at 2 mA. Dysarthria followed a similar pattern: absent at baseline and mild during 130 Hz tonic stimulation at 1 mA (rating = 2), but pronounced at 2 mA (rating = 4). Pink-noise stimulation at 2 mA resulted in complete resolution of speech impairment (rating = 0), while 3 mA caused only mild dysarthria (rating = 3).

#### Drawing-based tremor assessments

To more precisely characterize stimulation pattern effects on functional tremor control and side-effect profiles, spiral drawing tasks were performed across an expanded range of stimulation amplitudes using Program 2 contact configuration (Fig 5; Fig S3). Baseline drawings demonstrated significant tremor, while increasing stimulation amplitudes produced progressive improvement in spiral smoothness and Fahn–Tolosa–Marin (FTM) scores across both tonic and pink noise paradigms. Both stimulation types greatly reduced tremor relative to OFF; however, tonic stimulation was associated with increasing dysarthria and paresthesia scores at higher amplitudes, limiting the usable therapeutic window. In contrast, pink noise stimulation achieved comparable tremor suppression while maintaining lower side-effect burden across tested amplitudes.

#### Recharge and Noise Structure Comparisons

To determine whether observed differences could be explained by stimulation recharge characteristics rather than temporal structure, tonic 130 Hz stimulation was tested under both passive and active recharge configurations across matched amplitudes (Fig S4–S5). Across conditions, recharge mode produced similar tremor suppression and comparable side-effect profiles, suggesting that clinical differences observed between tonic and stochastic stimulation were unlikely to result solely from recharge dynamics. To isolate the effects of temporal structure, pink noise stimulation was compared directly with white noise stimulation using spiral and line drawing tasks (Fig 6; Fig S6). Both stochastic paradigms reduced tremor severity relative to OFF; however, pink noise demonstrated slightly lower FTM scores at intermediate amplitudes compared with white noise. Side-effect ratings remained low across stochastic conditions.

#### Home Trial Observations

Following the initial programming visit, the patient completed a blinded one-week home trial comparing tonic stimulation and two pink noise stimulation programs during routine daily activities. The patient reported slightly better tremor suppression with tonic stimulation but consistently noted clearer speech with both pink noise programs. Several coworkers independently commented that his speech sounded improved while using the pink noise settings. These observations suggested that stochastic stimulation preserved clinically meaningful tremor control while improving tolerability during everyday use and motivated subsequent in-clinic testing using higher amplitudes and additional temporal patterns, including white noise.

## Discussion

This first-in-human longitudinal case study demonstrates that stochastic pulse timing can preserve meaningful tremor suppression while reducing stimulation-induced dysarthria and paresthesias across repeated in-clinic and real-world testing. Across functional testing paradigms, pink noise–patterned stimulation achieved clinically meaningful tremor suppression without the dysarthria and paresthesias that emerged at comparable amplitudes with conventional tonic stimulation. These effects were reproducible across stimulation programs, amplitudes, test settings, and contact configurations, suggesting that temporal patterning represents a promising and clinically feasible programming dimension that could be implemented within standard DBS workflows.

High-frequency tonic stimulation has remained the standard DBS protocol for over two decades because of its robust suppression of motor symptoms and pathological network activity [1]. However, its highly regular temporal structure may also promote unintended entrainment of neural circuits, contributing to stimulation-induced side effects such as dysarthria and paresthesias [6]. These adverse effects are traditionally attributed to current spread or anatomical proximity to off-target structures, but our findings suggest that temporal regularity itself may play an independent role. In this study, tonic and pink noise stimulation were matched for mean frequency and amplitude, yet side effects emerged more prominently under tonic stimulation (Fig 4-5), implicating temporal organization rather than charge delivery alone as a contributing factor.

Temporally patterned stimulation introduces controlled variability into inter-pulse intervals while preserving overall charge delivery, enabling modulation of neural dynamics beyond conventional tonic stimulation [12, 13]. Pink noise–patterned stimulation incorporates structured, scale-free (1/f) temporal correlations that resemble statistical properties observed in many natural systems, whereas white noise introduces uncorrelated stochastic variability [14](Fig S2. In this study, both stochastic paradigms reduced side-effect burden relative to tonic stimulation, suggesting that temporal irregularity may disrupt rigid entrainment associated with adverse effects. However, pink noise demonstrated some improved tremor control compared to white noise (Fig 6) and was subjectively preferred by the patient in clinic and home settings, indicating that structured temporal organization—not randomness alone may be important.

The implications of temporally patterned stimulation extend beyond essential tremor and suggest a broader role for temporal structure as an independent programming dimension in neuromodulation. Many neurologic and psychiatric disorders treated with DBS involve pathological synchronization within distributed neural networks [1], and may also benefit from stimulation strategies that reduce entrainment and more closely approximate physiological dynamics. Adaptive DBS approaches have recently focused on optimizing when stimulation is delivered based on physiological biomarkers [17–19]. In contrast, our findings show that modifying how stimulation pulses are temporally structured—even when delivered continuously—may meaningfully influence both therapeutic efficacy and tolerability. Introducing structured temporal variability may allow disruption of pathological rhythms while avoiding excessive entrainment of adjacent circuits or off-target pathways. Because temporal patterning can be implemented through software-based control without changes to electrode placement or hardware design, this approach represents a potentially scalable strategy for expanding the therapeutic window across existing DBS platforms.

This study is limited by its single-subject, proof-of-concept design. The reproducibility of these findings across larger patient cohorts, additional anatomical targets, and longer-term follow-up remains to be established. Although pink and white noise paradigms were selected to represent structured and unstructured stochastic stimulation, other temporal architectures (e.g. other noise patterns, burst cycling) may offer different advantages and should be explored. Future studies incorporating larger cohorts, longitudinal assessment, and concurrent neurophysiological recordings will be necessary to define optimal temporal structures for specific neuromodulation targets and clinical indications.

## Conclusion

This study supports the feasibility and clinical utility of stochastic DBS pulse timing as an alternative to conventional tonic stimulation. By preserving tremor suppression while eliminating dysarthria and paresthesias, temporally structured variation in pulse timing may offer a path to expand the therapeutic window. Moreover, our findings suggest that the structure of variability—not just randomness—may be key to these benefits. Temporally patterned stimulation represents a promising direction for future neuromodulation strategies across neurologic and psychiatric disorders.

## Data Availability

All data produced in the present work are contained in the manuscript.

## Funding

This work was supported by the National Institutes of Health (NIH) NINDS U01-NS128612 (MRB, KJM). Manuscript contents are solely the responsibility of the authors and do not necessarily represent the official views of the NIH. MRB, KJM, and BTK were supported by the MN partnership grant for biotechnology and medical genomics (MNP2142). MRB and KJM were also supported by a Helene Houle Career Development Award and the Tianqiao & Chrissy Chen Institute. The funders had no role in study design, data collection and analysis, decision to publish, or preparation of the manuscript. ChatGPT (OpenAI) was used for assistance in text editing but not content generation. HB and SN are employees of Boston Scientific Corporation. Boston Scientific provided access to the Chronos research platform and technical/research support for this study but did not provide financial support to MRB, KJM, or BTK.

## Supporting information

**Fig. S1.**
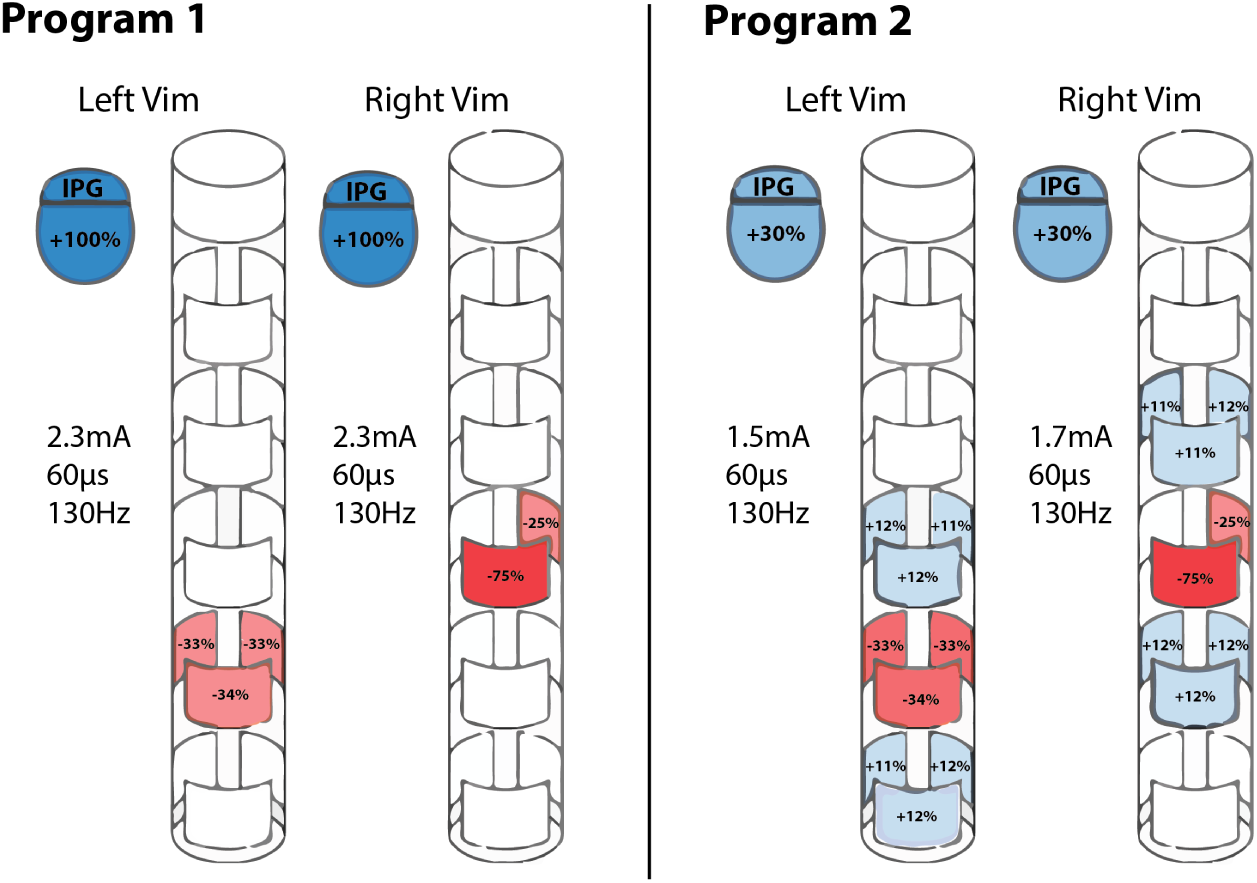
Patient Programming Contact Configurations. Programming configurations for both leads targeting the ventral intermediate nucleus (Vim). Program 1 utilized a monopolar configuration with cathodic stimulation distributed across a vertical contact triplet on each side and the IPG case serving as the anode (+100%). This setting was initially effective but resulted in stimulation-induced dysarthria. Program 2 employed a bipolar configuration with a more distributed current spread across multiple directional contacts and the anode set within the lead (+30% IPG). This configuration reduced dysarthria and became the patient’s clinical program at the start of the study, though mild speech effects persisted.

**Fig. S2.**
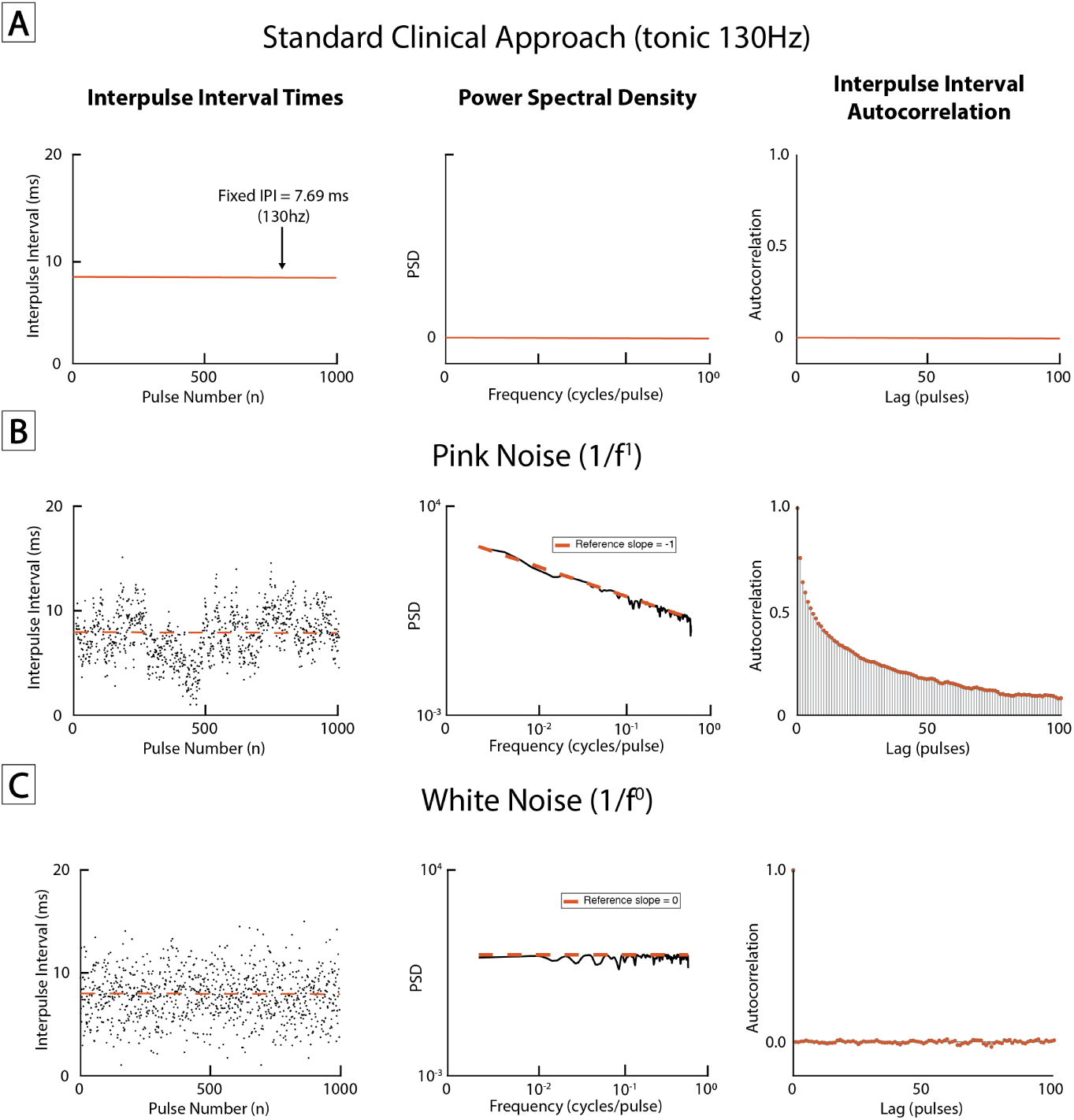
Representative temporal structure of noise-colored stimulation pulse trains. **(A)** Standard clinical tonic stimulation (fixed 130 Hz), demonstrating constant interpulse intervals (IPI), a flat 0 power spectral density (PSD), and no temporal autocorrelation beyond lag 0. **(B)** Pink noise–patterned stimulation (1*/f* ^1^), showing stochastic variation in interpulse intervals with a power-law PSD slope and long-range temporal correlations reflected in the autocorrelation function. **(C)** White noise–patterned stimulation (1*/f* ^0^), demonstrating randomly varying interpulse intervals with a flat PSD and no temporal autocorrelation. Left column: interpulse interval time series across pulses. Middle column: power spectral density of the interpulse interval sequence. Right column: autocorrelation of interpulse intervals across pulse lags.

**Fig. S3.**
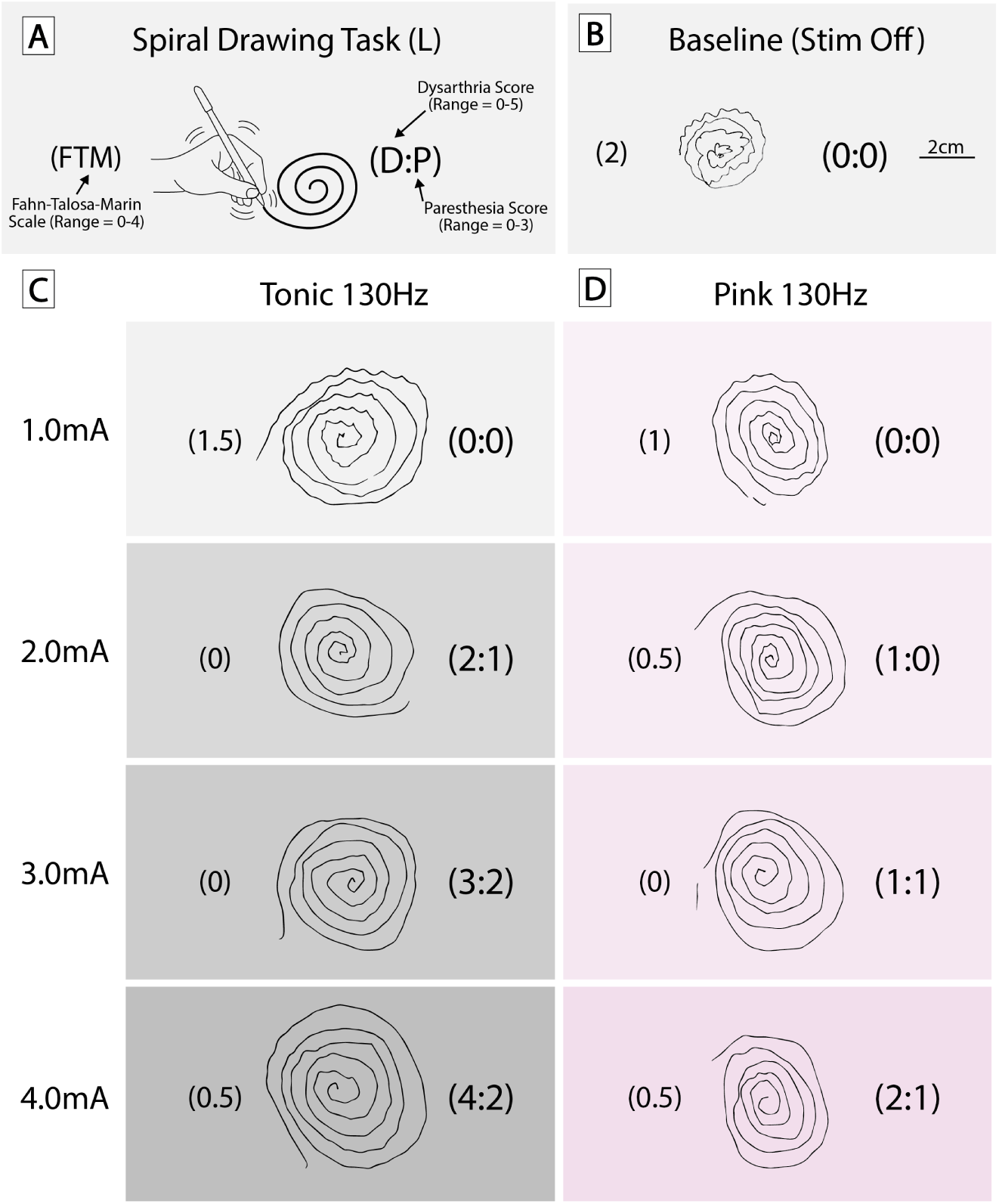
Left-hand spiral drawings and side effect profiles during in-clinic stimulation testing comparing tonic and pink noise stimulation. **(A)** Spiral drawing task schematic illustrating the Fahn–Tolosa–Marin (FTM) tremor scale (0–4), dysarthria score (0–5), and paresthesia score (0–3). **(B)** Baseline spiral drawing with stimulation OFF. Right-hand spiral drawings obtained during in-clinic testing under tonic 130 Hz **(C)** and pink noise–patterned 130 Hz **(D)** stimulation at 1.0, 2.0, 3.0, and 4.0 mA. FTM tremor scores are shown left of each spiral, and dysarthria and paresthesia scores are displayed in parentheses to the right (dysarthria:paresthesia). Both stimulation paradigms reduced tremor relative to OFF across amplitudes. At higher amplitudes, tonic 130 Hz was associated with increasing dysarthria and/or paresthesia, whereas pink 130 Hz achieved comparable tremor suppression with lower side effect scores. All stimulation conditions were tested using program 2 contact configurations (Fig S1).

**Fig. S4.**
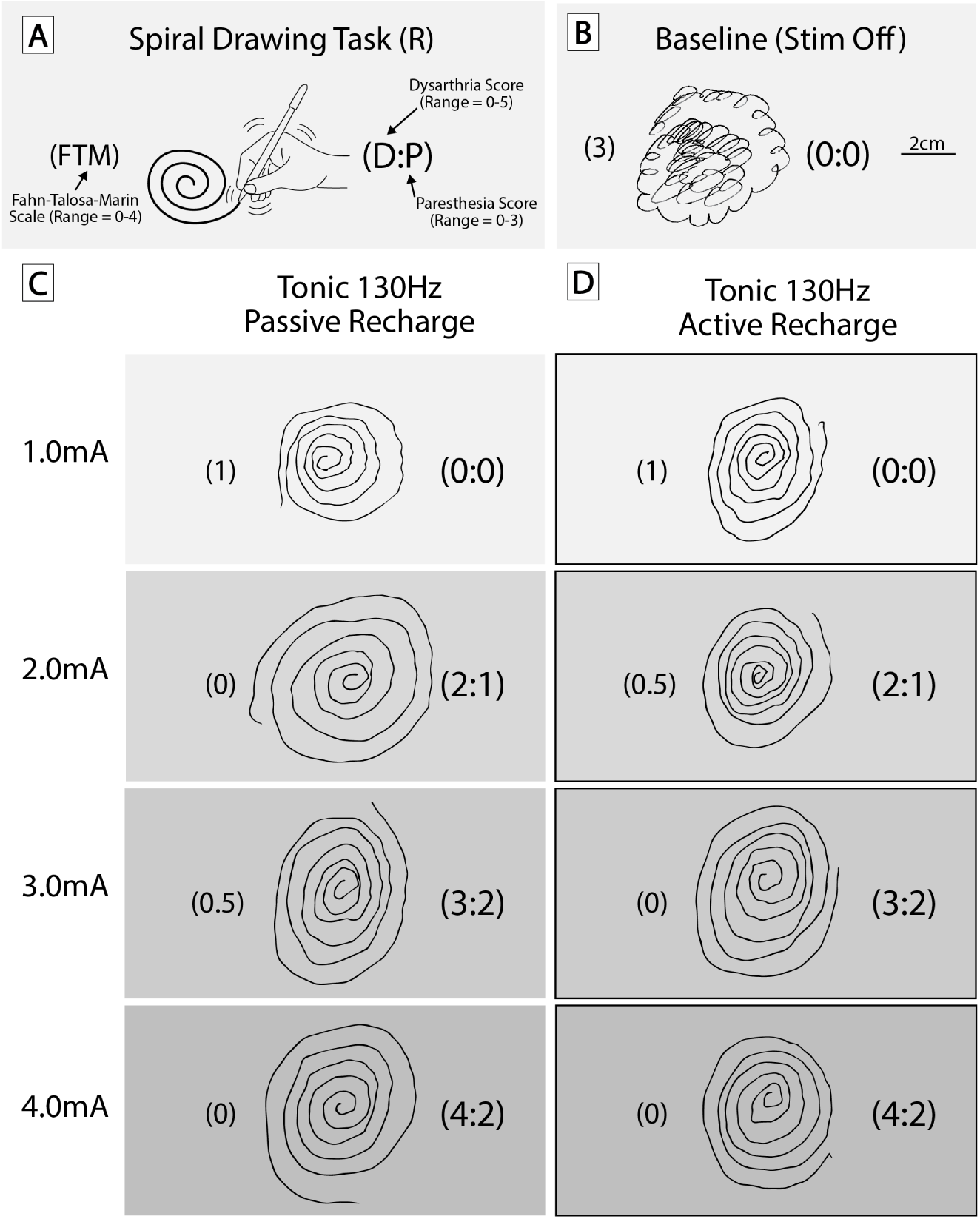
Right-hand spiral drawings and side effect profiles during in-clinic stimulation testing comparing active and passive recharge for tonic stimulation. **(A)** Spiral drawing task schematic illustrating the Fahn–Tolosa–Marin (FTM) tremor scale (0–4), dysarthria score (0–5), and paresthesia score (0–3). **(B)** Baseline spiral drawing with stimulation OFF. Right-hand spiral drawings obtained during in-clinic testing under tonic 130 Hz **(C)** and pink noise–patterned 130 Hz **(D)** stimulation at 1.0, 2.0, 3.0, and 4.0 mA. FTM tremor scores are shown left of each spiral, and dysarthria and paresthesia scores are displayed in parentheses to the right (dysarthria:paresthesia). Both recharge configurations reduced tremor severity relative to OFF across amplitudes and showed similar side effect profiles with increasing amplitudes. All stimulation conditions were tested using program 2 contact configurations (Fig S1).

**Fig. S5.**
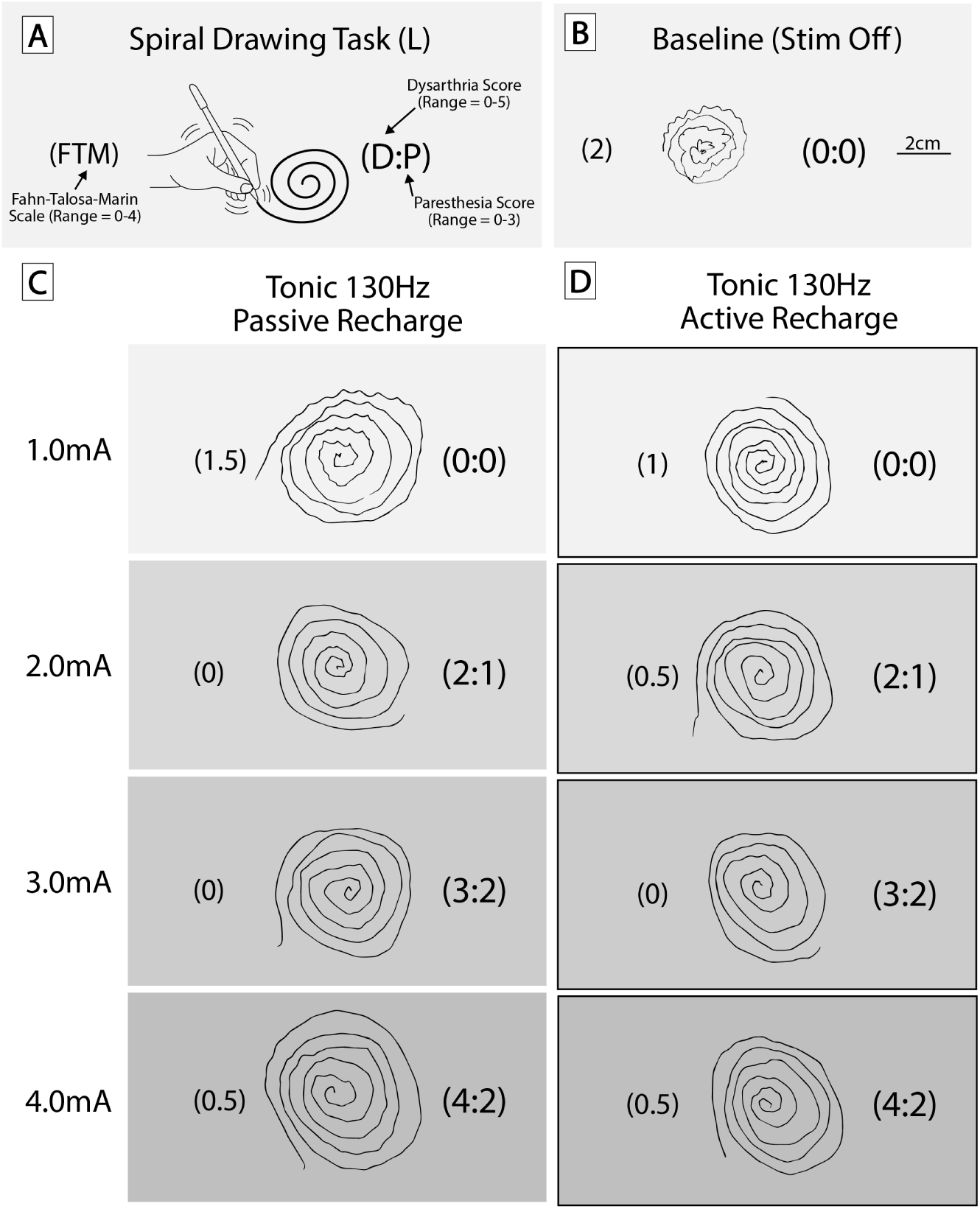
Left-hand spiral drawings and side effect profiles during in-clinic stimulation testing comparing active and passive recharge for tonic stimulation. **(A)** Spiral drawing task schematic illustrating the Fahn–Tolosa–Marin (FTM) tremor scale (0–4), dysarthria score (0–5), and paresthesia score (0–3). **(B)** Baseline spiral drawing with stimulation OFF. Right-hand spiral drawings obtained during in-clinic testing under tonic 130 Hz **(C)** and pink noise–patterned 130 Hz **(D)** stimulation at 1.0, 2.0, 3.0, and 4.0 mA. FTM tremor scores are shown left of each spiral, and dysarthria and paresthesia scores are displayed in parentheses to the right (dysarthria:paresthesia). Both recharge configurations reduced tremor severity relative to OFF across amplitudes and showed similar side effect profiles with increasing amplitudes. All stimulation conditions were tested using program 2 contact configurations (Fig S1).

**Fig. S6.**
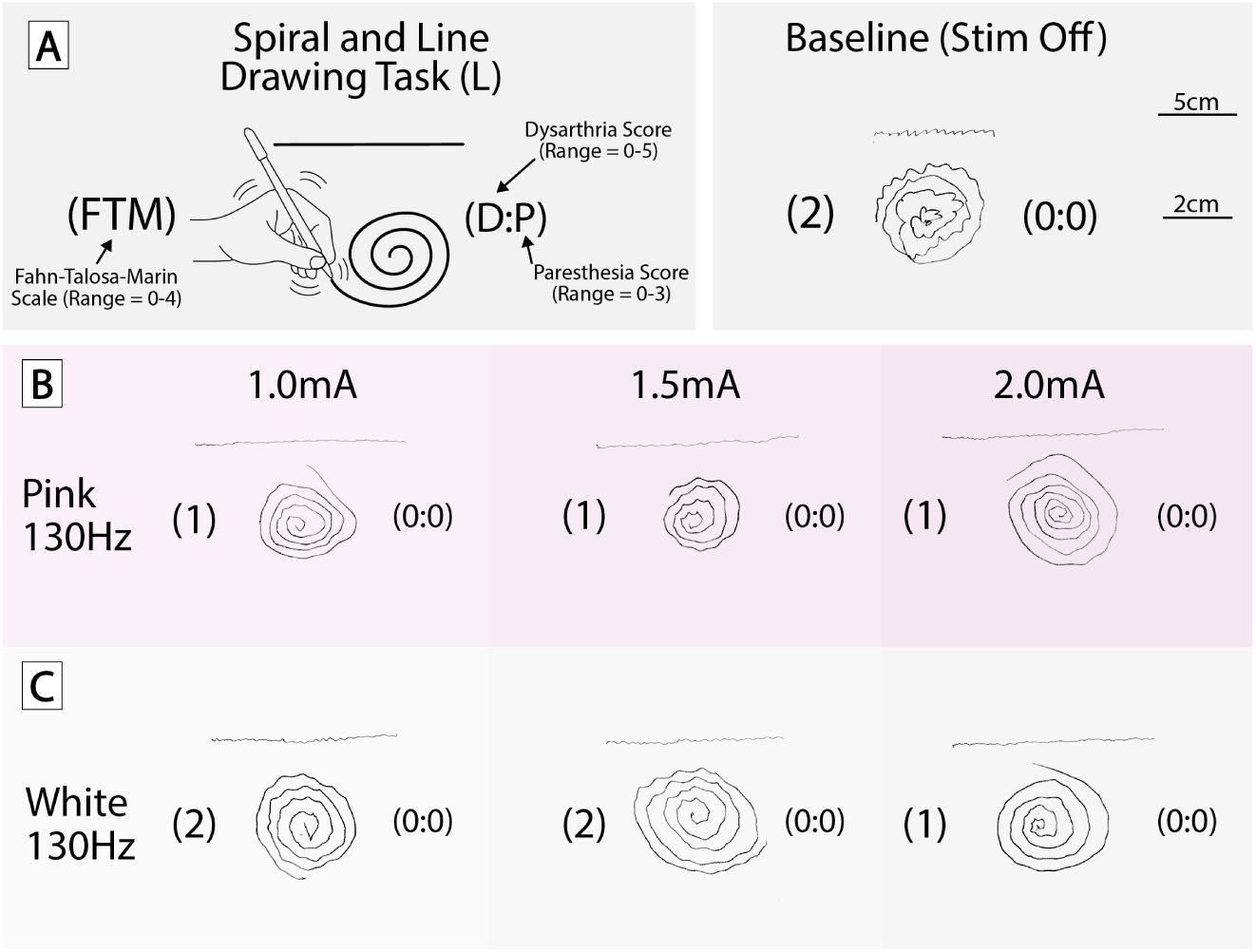
Left-hand spiral and line drawings during in-clinic stimulation testing comparing pink and white noise stimulation. **(A)** Spiral and line drawing task schematic illustrating the Fahn–Tolosa–Marin (FTM) tremor scale (0–4), incorporating both spiral and line components, along with dysarthria score (0–5) and paresthesia score (0–3). **(B)** Baseline drawing with stimulation OFF. Representative right-hand drawings obtained during in-clinic testing under pink noise–patterned 130 Hz **(C)** and white noise–patterned 130 Hz **(D)** stimulation at 1.0, 1.5, and 2.0 mA. FTM tremor scores are shown to the left of each drawing, and dysarthria and paresthesia scores are displayed in parentheses to the right (dysarthria:paresthesia). Both stimulation paradigms reduced tremor severity relative to OFF, with pink noise stimulation demonstrating slightly lower FTM scores at intermediate amplitudes compared with white noise. All stimulation conditions were tested using program 2 contact configurations (Fig S1).

